# Time Dynamics of COVID-19

**DOI:** 10.1101/2020.05.21.20109405

**Authors:** Cody Carroll, Satarupa Bhattacharjee, Yaqing Chen, Paromita Dubey, Jianing Fan, Álvaro Gajardo, Xiner Zhou, Hans-Georg Müller, Jane-Ling Wang

**Affiliations:** Department of Statistics, University of California, Davis, Davis, CA 95616 USA

**Keywords:** COVID-19, Cases and Deaths, Doubling Rates, Case Fatality Rates,, Functional Data Analysis, Functional Principal Component Analysis, Empirical Dynamics, Concurrent Regression

## Abstract

We apply tools from functional data analysis to model cumulative trajectories of COVID-19 cases across countries, establishing a framework for quantifying and comparing cases and deaths across countries longitudinally. It emerges that a country’s trajectory during an initial first month “priming period” largely determines how the situation unfolds subsequently. We also propose a method for forecasting case counts, which takes advantage of the common, latent information in the entire sample of curves, instead of just the history of a single country. Our framework facilitates to quantify the effects of demographic covariates and social mobility on doubling rates and case fatality rates through a time-varying regression model. Decreased workplace mobility is associated with lower doubling rates with a roughly two week delay, and case fatality rates exhibit a positive feedback pattern.

## 1 Introduction

As of May 1, 2020, more than 3 million cases of COVID-19 have been reported worldwide, leading to more than 200, 000 coronavirus related deaths [1]. The World Health Organization declared the situation a pandemic on March 11, 2020, and nearly all countries have been exposed to SARS-CoV-2, the betacoronavirus which causes the disease [2]. Despite the far reach of the virus, the pattern and rate of its spread within a population is not uniform. Some countries like the US have seen marked increases in case and death counts per capita, even after implementing distancing measures, while others like Japan have been able to keep the spread of disease low for long durations despite comparatively lax social restrictions. Measures that mitigate spread in one case may not work uniformly across countries due to effects of demographics and timing, among other factors. Data-driven analyses of the time-dynamics of cases and deaths are of central importance to characterize underlying forces and unexplained variation.

Global efforts to “flatten the curve” of COVID-19 cases translate quantitatively to decreasing epidemiological statistics like doubling rates via social distancing campaigns, improved hygiene and case tracking. Early statistical inquiries have focused on estimation of doubling rates and case fatality rates with SIRD and SEIM models [3, 4] and forecasting the number of cases worldwide using time series analysis [5]. The analysis of the effects of prevention efforts like social distancing for specific countries has also received increasing attention [6, 7].

Processes that grow exponentially, such as the case load of unmitigated COVID-19 transmissions are characterized by a fixed doubling time. In reality, however, the doubling time is a dynamic quantity, which changes every day due to mitigation efforts and the changing inherent dynamics of virus spread. It is then vital that policy makers have access to frequent and up-to-date estimates of doubling time [8]. In recent work [9, 10] the basic reproduction number and doubling time have been studied in a dynamic manner by considering a varying coefficient model with daily new cases as the response and time as a predictor. Similar work has considered the real-time estimation of case fatality rate using Poisson mixture models [11]. Our analysis complements these studies and we propose an alternative way of obtaining relevant dynamic quantities. Associating metrics of disease progression with baseline covariates is also of central interest. Recent research has queried the effects of population age, temperature, humidity [12, 13], community mobility patterns [14, 15] and other factors on the spread of the virus. Modeling the transmission dynamics of COVID-19 based on predictors is key to understanding the impact of distancing practices and other policies on spread mitigation and prevention. We refer to [16] for a more thorough review of recent epidemiological analyses.

It should be noted that COVID-19 analyses based on published case and death counts, including those conducted here, are subject to the same biases which affect the accuracy of the data, primarily due to under-reporting [17], the degree of which varies by country [18]. The reasons for such under-reporting are many, including insufficient testing materials, political incentives, and administrative delays. With this caveat in mind, the study of available data may nevertheless provide useful insights and stimulation for further research.

We propose here functional data analysis as a tool for analyzing the time-dynamics of COVID-19 as quantified by case and death numbers across countries. Functional data analysis (FDA) [19, 20, 21] aims to detect patterns and quantify samples of random trajectories through functional principal component analysis [22, 23], empirical dynamics [24] and other methods where entire curves are viewed as data atoms. This methodology is uniquely suited for the analysis of COVID-19 data since the cumulative case counts across countries amount to a sample of random curves observed over time.

The cumulative COVID-19 case and death count trajectories per million people (in log scale) for 64 countries are shown in Figure 1 (see Methods for further details). The trajectories are shown for a 67-day interval after the first time a country reports at least 20 confirmed cases which is defined as the origin of the time scale and thus corresponds to different calendar times (see Data).

**Figure 1:**
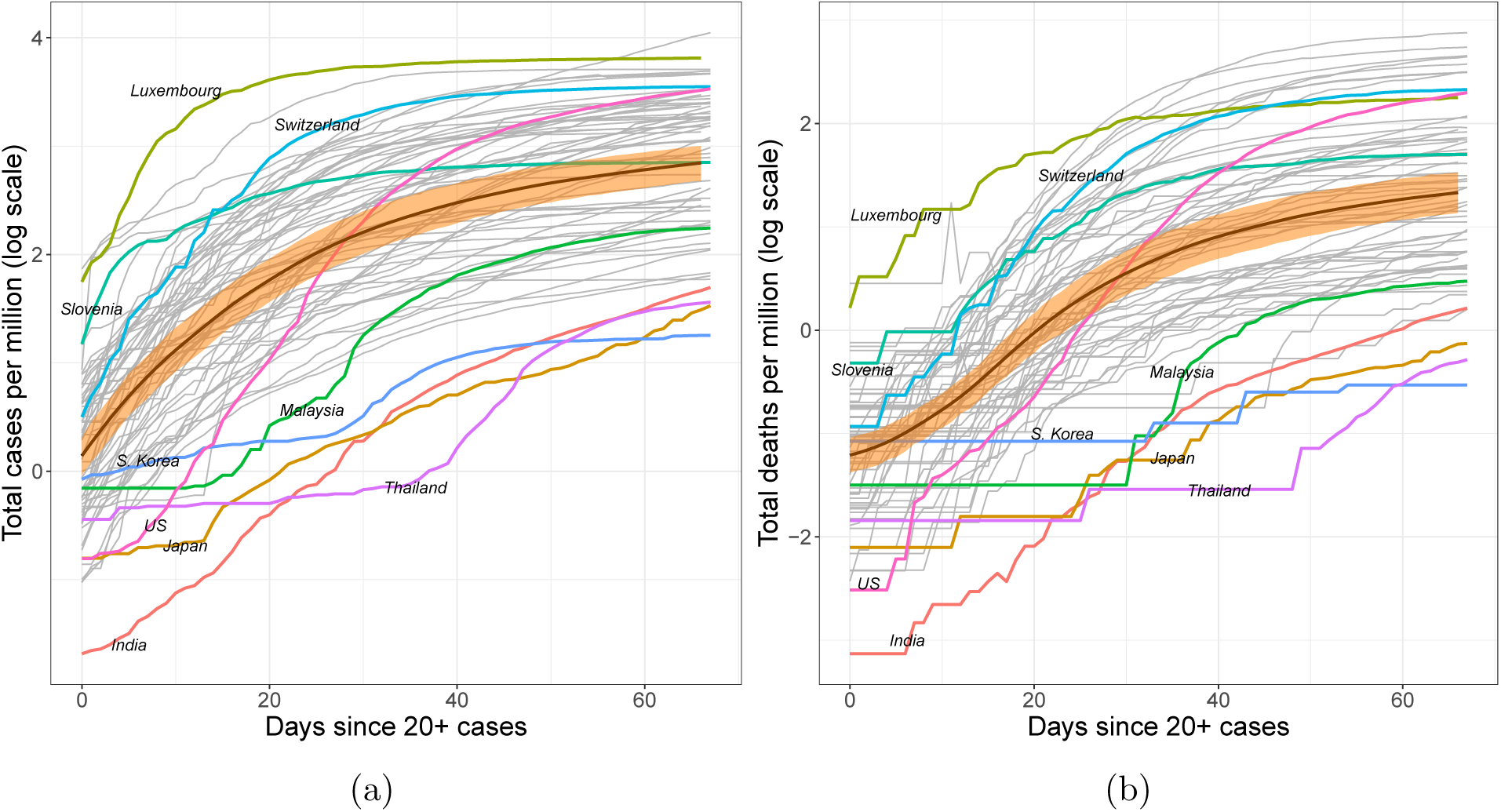
Trajectories of (a) total case count and (b) total death count per million individuals on log scale. The time window spans the 67 days since at least 20 confirmed cases were reported. Smoothed mean curves are marked by bold black lines. The orange ribbons represent pointwise 95% confidence bands for the overall mean functions.

Functional approaches are uniquely suited for the modeling, interpreting, and predicting trajectories of cumulative case counts, as they leverage the latent information shared between countries to boost the efficiency of predictions and facilitate comparisons of the trajectories across countries. We use FDA methodology to study patterns of growth for case counts and to quantify the performance of various countries as the pandemic progresses. We also showcase an approach for predicting future case counts which may be used to understand whether a country is doing better or worse than expected. Finally, we explore the role of variables such as population density, fraction of the population over 65 and mobility reduction efforts for the spread of the infection as it varies over time. Throughout, our focus is on associations which may suggest but do not establish causality.

## 2 Results

### 2.1 COVID-19 dynamics across countries

#### 2.1.1 Main patterns of disease propagation

For the 64 countries in the study, case counts per million generally follow one of four paths over time. They are either 1. consistently higher than average (e.g., Switzerland), 2. consistently lower than average (e.g., India), 3. initially lower but then experience a dramatic increase over time (e.g., the US), or 4. initially higher before entering a period of control (e.g. Slovenia). These archetypes are derived from the extreme ends of the two main modes of variation [25, 26] observed for the sample, which emerge from functional principal component analysis (FPCA, Figure 2). FPCA is similar to ordinary principal component analysis in the sense that it projects high dimensional curve data into a low dimensional space, representing them as a random vector of functional principal component (FPC) scores (see Methods). These scores are matched with smooth eigen*functions*, which reveal the primary patterns of variation in the sample of functional objects. For the case load trajectories, a 2-dimensional representation was found to adequately represent the sample of cumulative case trajectories.

**Figure 2:**
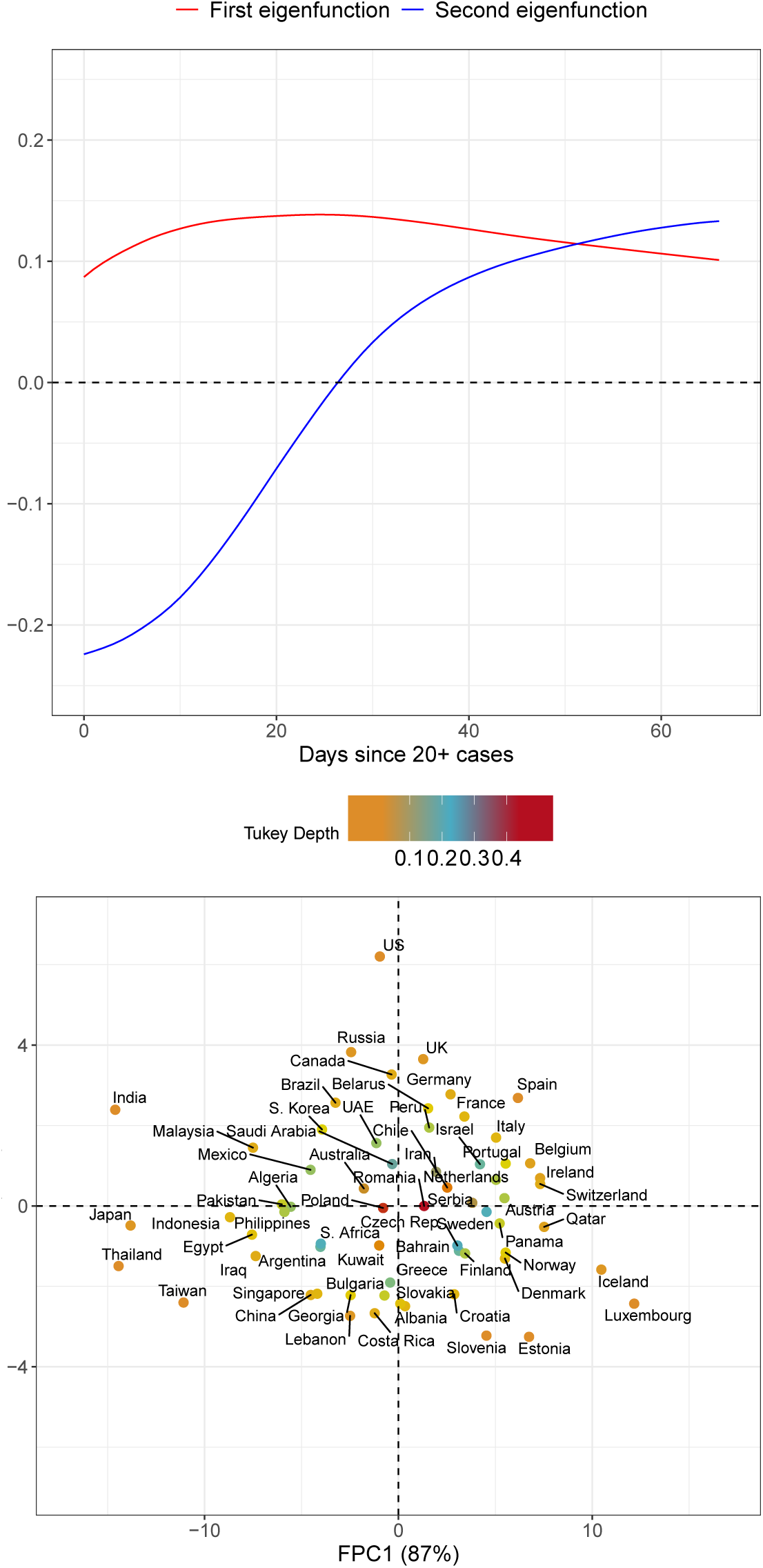
Visualizing (a) the first (red) and second (blue) eigenfunctions and (b) their corresponding FPC scores. The two dimensional representation in FPC space captures 97% of the variation in sample of case load trajectories across countries. Points are colored according to their Tukey depth, where higher depths indicate trajectories closer to the mean curve.

An inspection of the eigenfunctions and their corresponding FPC scores reveals which of the four patterns a country generally follows (Figure 2). FPC scores of countries with consistently higher case trajectories are located in the right half-plane, and those with uniformly lower rates in the left. Analogously, countries which experience a dramatic increase in cases per million lie in the upper half of the plane, and those which successfully slow their spread in the lower half. Countries which follow similar trajectories are clustered together in FPC space, with outliers located in the outskirts of the point cloud. While some outliers are apparent from a simple visual inspection of the original curves (e.g. Luxembourg, Thailand, India and Japan), other atypically-shaped trajectories are initially obscured in the crowd of curves until revealed in FPC space (e.g. the US).

The concept of modes of variation is useful for visualizing the range of FPC scores as a spectrum of curves (Figure 3). For example, India’s deviation from the mean curve is largely explained by just the first mode of variation. Its very negative first FPC score places it far below the mean throughout the entire interval. In this sense the first FPC score is similar to a random intercept in a linear mixed model, since the first eigenfunction is roughly constant over time. Incidentally, the slight curvature exhibited in this eigenfunction allows for flexible modeling of the “curve bending” phenomenon as demonstrated by Switzerland, which has a very positive first FPC score.

**Figure 3:**
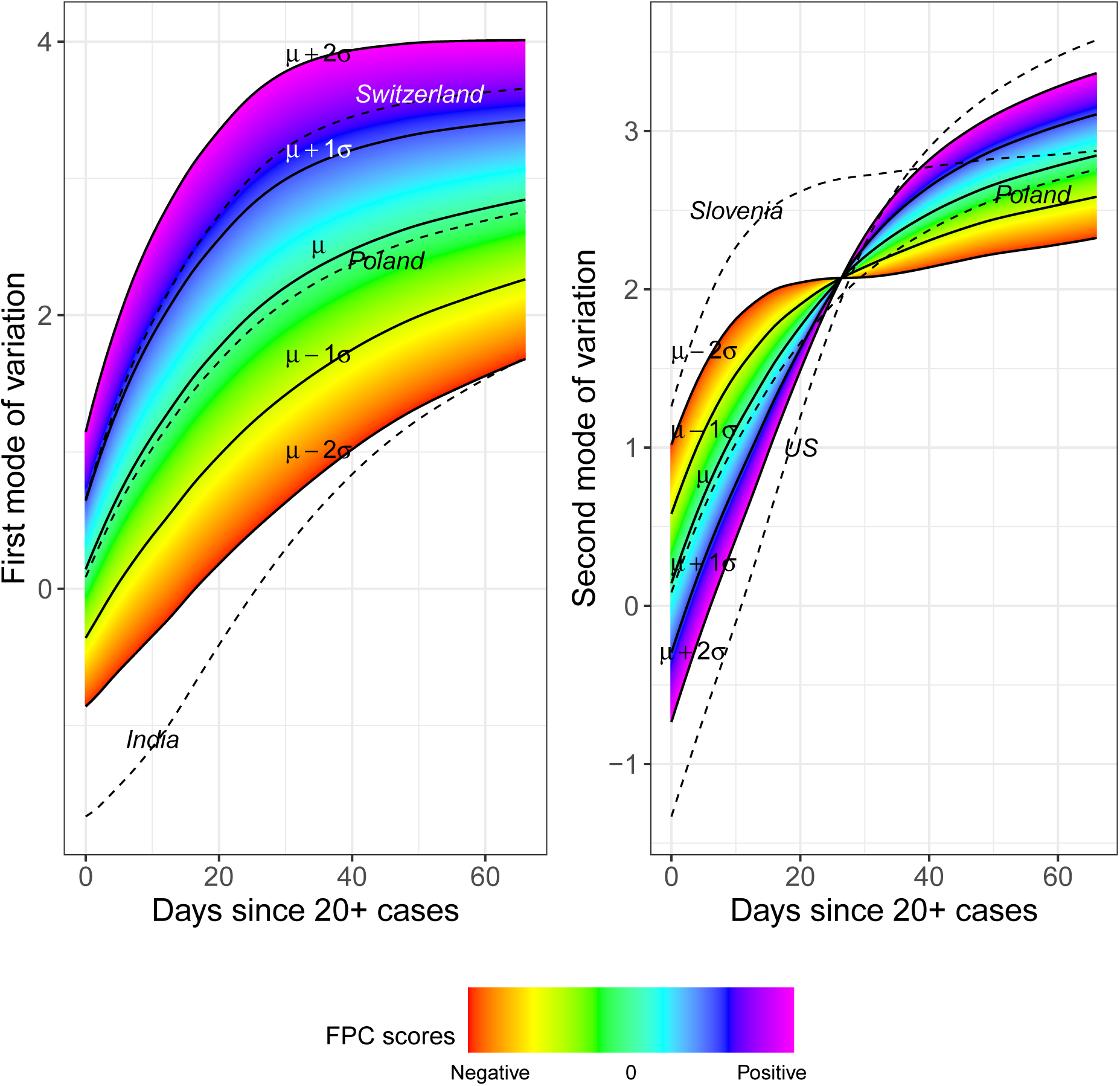
First and second modes of variation for the total cases process, where the sample standard deviations of the first and second FPC scores are 5.77 and 1.96 respectively. The dashed lines correspond to the fitted trajectories for some countries.

The US departs from the typical mean trajectory in a different way: Its case count per capita has increased dramatically by the end of the time interval, which is seen by its very positive second FPC score. The second mode of variation captures a subtle curvature that would be missed in parametric modeling. In this sense the second FPC score modulates the shape of the curve rather than its magnitude. Nearly all cumulative case trajectories begin to level off after roughly *t* = 25 days, a trend which is illustrated neatly by the inflection point in the second mode of variation. One may then interpret the first month of exposure as a sort of “priming period,” after which (non-outlier) trajectories exhibit more stable trends. This interpretation echoes the findings of [27], who also identify the first few weeks of exposure as a critical time for disease management.

Countries which are clustered together in FPC space may also share other underlying structures such as geographic proximity, exemplified by the Western European countries in the upper-right quadrant of Figure 2(b). In some cases, far-flung places may act as if they are part of a distinct geographic cluster, such as Panama and Qatar with trajectories that are very similar to those in the Scandinavian block.

Countries’ performances in the two main modes of variation can be evaluated by comparing their respective FPC scores. Higher scores indicate higher rates of spread for both modes of variation. For example, Switzerland has a much higher first FPC score than India, reflecting that the case count per capita is higher for Switzerland throughout the time period. Similarly, the case trajectory in the US is seen to have shot up much more over time than that of Slovenia, say, since the second FPC score of the US is very large and positive while it is negative for Slovenia.

#### 2.1.2 Comparisons via rank dynamics

While FPC scores are useful for comparing and classifying country trajectories, they require multivariate comparisons, since each country has two scores. It is useful to complement these comparisons by ranking each country by cumulative cases counts per million, where higher percentile ranks correspond to increased infection rates and a generally worse situation. Such rankings can be done at each fixed time and then analyzed with rank dynamics [28]. The percentile ranks and their time-evolutions are illustrated in Figure 4 with a few notable curves highlighted. Higher percentiles signify more cases per capita. Switzerland’s transmission rates have been among the most severe, while India has performed well consistently throughout the time period. Both countries’ integrated ranks are among the highest and lowest, respectively, where integrated rank is the average rank over time. Both display low rank volatility (see Section S.6 in the Supplement), which is a measure of how ranks change over time, as their overall positions remain relatively stable throughout the time period.

**Figure 4:**
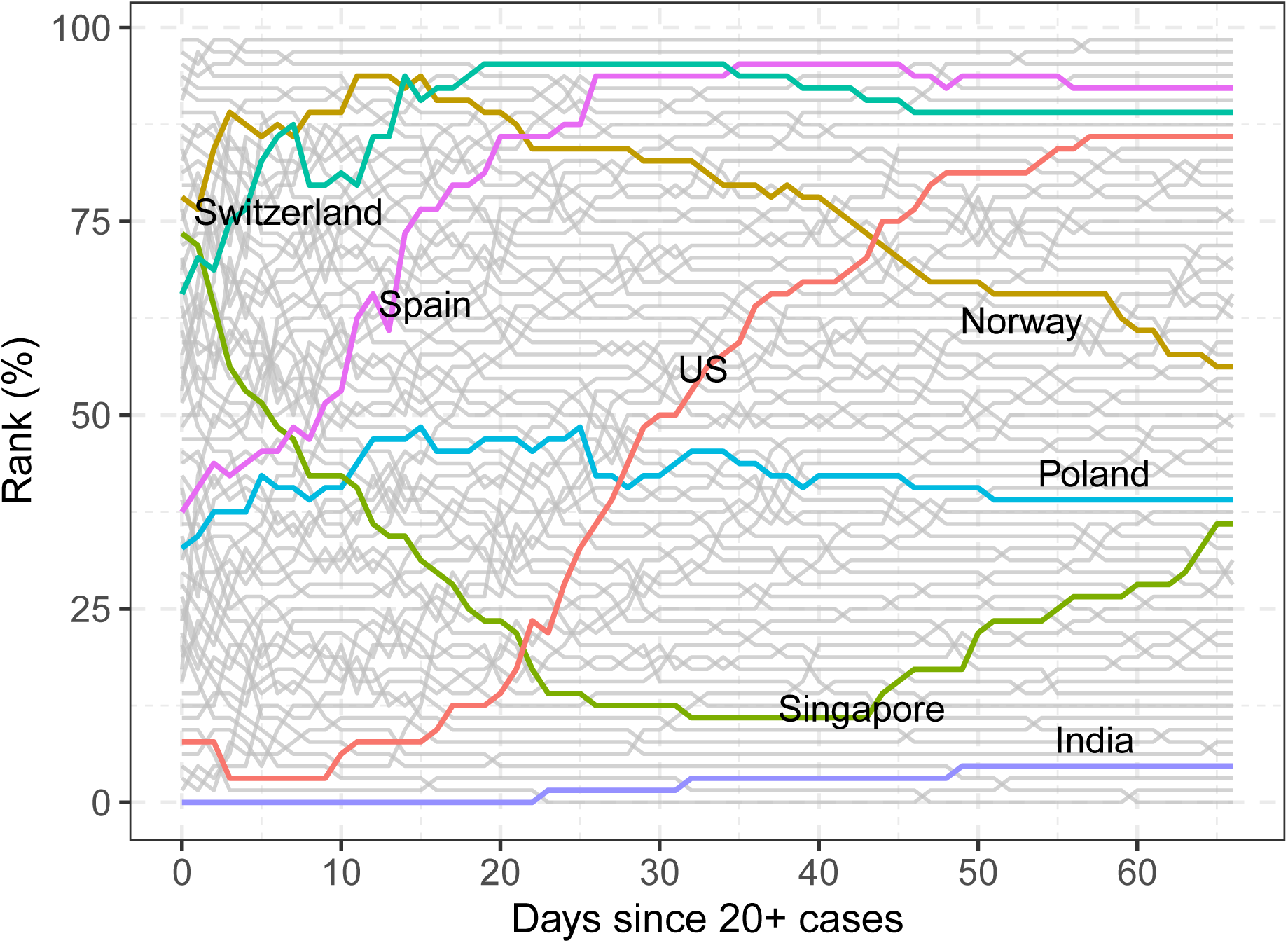
Rank trajectories during the first 67 days since exposure.

In terms of ranks, the situations in Spain and the US have substantially worsened over the time period, in line with the FPCA results, where Spain and the US were found to have large second FPC scores with associated strong increasing trends. These large shifts are reflected in their rank volatility, where Spain and especially the US visibly stand out (Section S.6 in the Supplement). On the other hand, the epidemic situations in Norway and Singapore have improved over time relative to other countries, especially during the first 40 days for the latter. However, Norway still has a somewhat severe situation overall, and Singapore’s percentile rank starts to rise in the last one third of the period. In contrast to these extremes, Poland has relatively stable ranks over time and also moderate epidemic situations compared to other countries. Overall the percentile trajectories are more volatile during the first 25 days. By the end of the interval however, the ranks have become relatively stable, as seen by the fewer number of intersections between paths.

### 2.2 Time evolution and forecasting of case trajectories

#### 2.2.1 Frequently-updating forecasts for viral transmission

Since the virus reaches countries at staggered times, countries with earlier exposure times hold valuable information for predicting those which were more recently exposed. This makes it possible to employ a dynamic FPCA approach to make short-term predictions of future trajectories by borrowing “future” information from countries for which a trajectory over a longer time interval is available, due to their earlier exposure in calendar time. Like FPCA, this is based on the assumption that there are common shape features that explain much of the variation. Borrowing of information from trajectories observed over a longer time stretch to inform trajectory prediction for those observed over a shorter time period is the key for dynamic FPCA.

The dynamic FPCA (dynFPCA) approach aims at continuously predicting future trajectories based on the most recently available data. Motivated by the rapidly updating nature of the COVID-19 case trajectories and their FPCA representation, dynFPCA harnesses the evolving trajectory data to predict future cumulative case counts. We illustrate this for 10-day-ahead predictions that are constructed on repeating intervals. Then, after the 10-day forecast has passed, one can compare the actually observed trajectories with the predicted trajectories over this interval. If the prediction underestimates the number of cases per capita this means that the country experienced a higher caseload than expected which could be due to a worse than average performance or to increased testing. Similarly an over-estimate in the prediction signifies a better than expected performance at the time of the prediction.

Since FPCA summarizes an observed curve as a finite vector of FPC scores, it is enough to predict just the FPC scores for the curve we wish to forecast. The scores may then be translated back into a curve on the entire interval, even if the scores are only estimated using a partially observed trajectory (see Methods: dynFPCA). The FPC scores are estimated using the conditional expectation approach [29], and the number of scores to estimate was chosen by an AIC criterion. The expression for conditional expectation contains quantities which are estimated from the entire sample of curves and thus the predictions borrow strength from the data of all countries. See the discussion in Methods for more technical details.

The predictions can be viewed in conjunction with the reduction in workplace mobility per country as reported by Google COVID-19 Community Mobility Reports [30] to quantify the magnitude of voluntary or mandated lock-downs (see Methods: Data). Early studies of the effects of lock-downs have estimated the delay until they affect the caseload to be roughly two weeks, roughly in line with the delay between infection and a recorded positive test result [6, 31].

Japan is a visible outlier in both measures (Figure 5), in the sense that it has a very low and flat cumulative case curve, while mobility levels do not decrease much, as the Japanese government did not issue lock-down orders and employers by and large continued to require physical presence at the workplace. Other countries with relatively flat curves include India and the Philippines. These countries acted early, as evidenced by major drops in workplace mobility early in the trajectory when the total caseload was still low.

**Figure 5:**
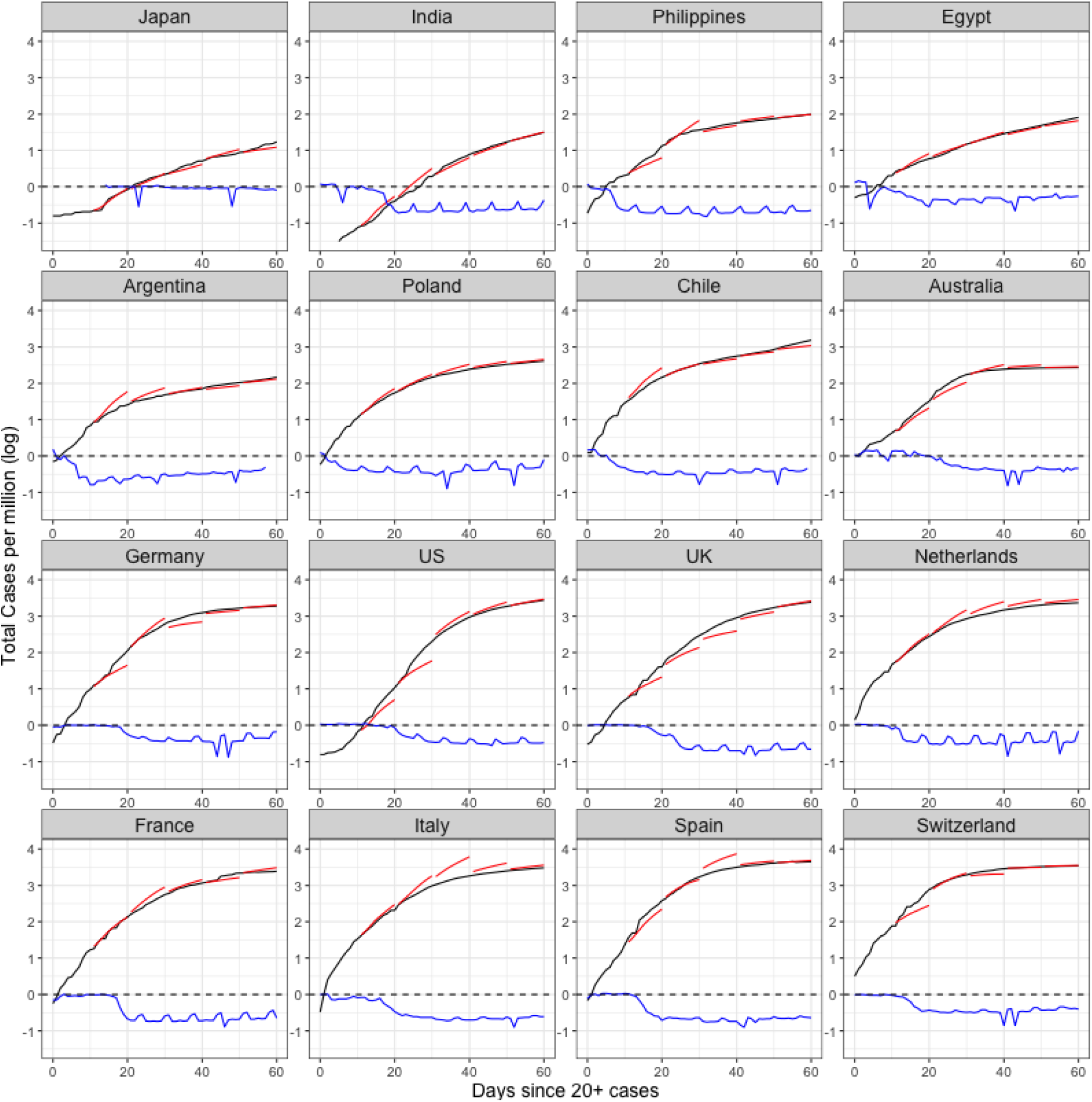
Forecasts of case counts for countries with early (top 8) and late (bottom 8) mobility reduction responses. The black curve represents the observed cumulative case counts, and the red piece-wise curve represents prediction based on first 10, 20, 30 and 40 days, respectively. The blue curve denotes percent decrease in workplace mobility as measured by Google.

Another group of countries with early drops in mobility includes Argentina, Chile and some other countries in South America as well as Poland. For countries that quickly reduced mobility, dynFPCA forecasts (and the eventually observed counts) reach much lower heights than those which did not. On the other hand, a group of mostly European countries (including Germany, the UK, the Netherlands, France, Italy, Spain and Switzerland) showed reduced mobility only after there were already a significant number of cases present in the population, which appears to have been too late to slow the momentum of infectious spread. As a consequence, their trajectories not only increase more rapidly but also reach higher overall levels than predicted. After the first two 10-day periods, which is characterized by high volatility as seen in the rank analysis, the dynFPCA method tends to predict the trajectories very accurately once curves begin to bend flatter.

#### 2.2.2 Time-dynamics for correlates of doubling rates and case fatality

To quantify the effect of mobility reduction on dynamic FPCA predictions, we studied its effect on doubling rate, in addition to the effects of other demographic predictors that might play a role in shaping the trajectories. Understanding the factors which correlate with increased or decreased rates of infection is critically important for policymakers and societies. As local situations evolve and the pressure to reopen mounts, an equally important aspect is understanding how these associations might change over time, and when predictors are particularly relevant, as it is likely that the effect of predictors is not stationary but rather varies over the course of the infection.

To investigate the time-varying relationship between a country’s demographics and mobility reduction and outcome measures like the doubling and case fatality rates, we applied a functional concurrent regression model and empirical dynamics (see Methods). Doubling rate, *γ*(*t*) and case fatality rate, CFR(*t*), as time-varying responses have been traditionally used in epidemiological modeling [16, 27, 32, 33]. Low doubling rates indicate successful containment of viral transmissions, while low case fatality rates indicate better outcomes in terms of the mortality of infected populations. Mean doubling rate and case fatality rates over time are depicted in Figure 6. We caution that estimates of the case fatality rate contain additional uncertainty, since the virus’ asymptomatic rate is not currently well understood.

**Figure 6:**
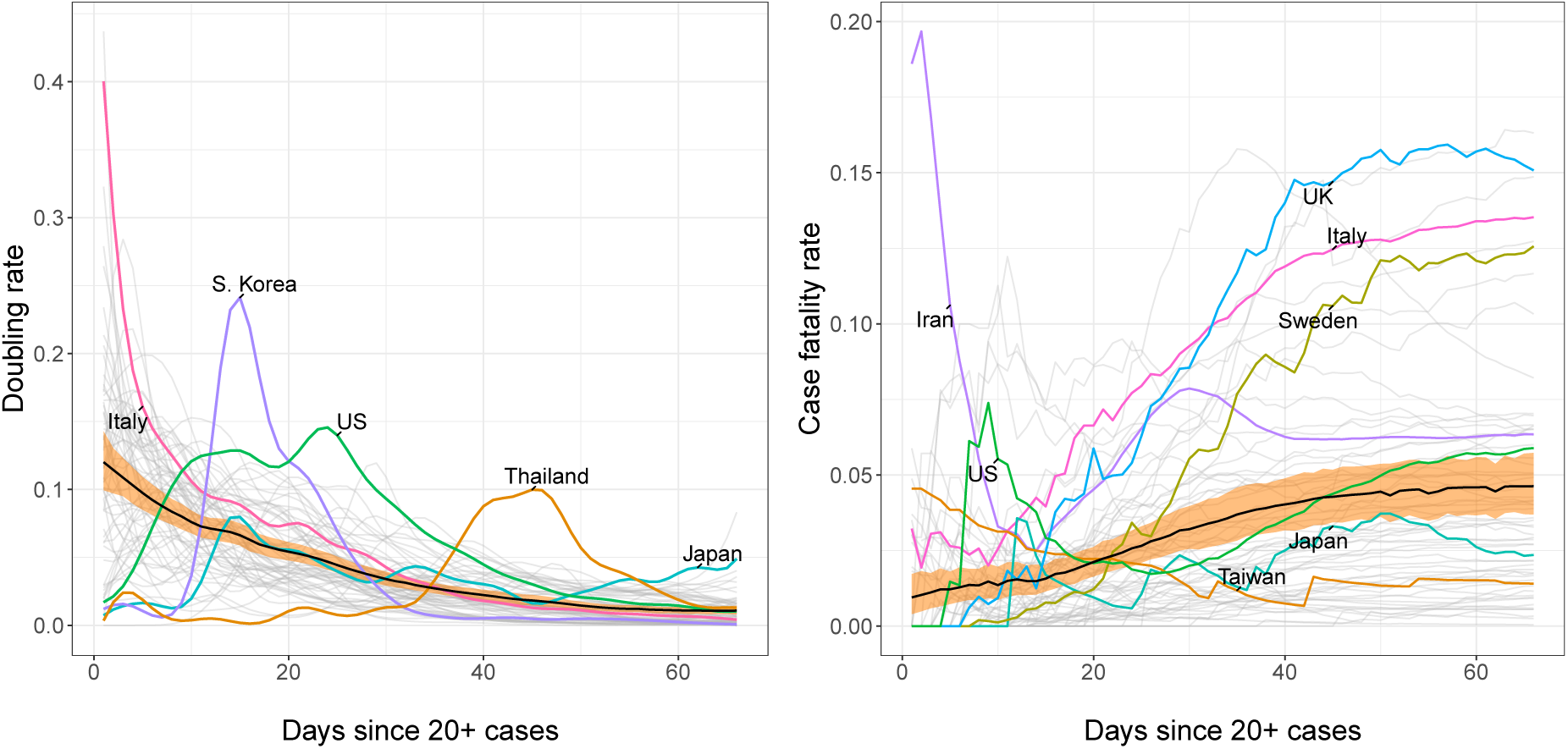
Time varying mean doubling rate (left) and mean case fatality rate (right) across all countries. The orange ribbon represents pointwise 95% confidence bands for the overall mean function.

We adopt the functional concurrent regression (FCR) models,

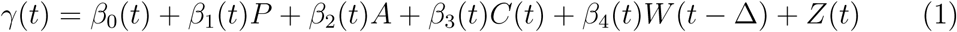

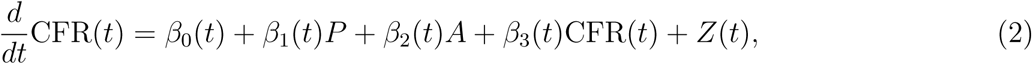

where the predictors consist of population density *P*, the proportion of the population over age 65 A, the log-cumulative case counts per million *C*(*t*), and the lagged decrease in workplace mobility *W* (*t* – Δ). The former two predictors are baseline covariates while the latter two are time-varying. The error term *Z* (*t*) denotes a mean-zero stochastic drift process. The optimal lag Δ is chosen data-adaptively and was found to maximize predictive power at Δ = 13 days (Methods: Empirical Dynamics). Concurrent effect functions for each regressor are displayed alongside their 95% confidence bands in Figure 7. A stretch of time where the confidence band does not touch 0 suggests that the effect of that predictor is locally significant during that time interval.

**Figure 7:**
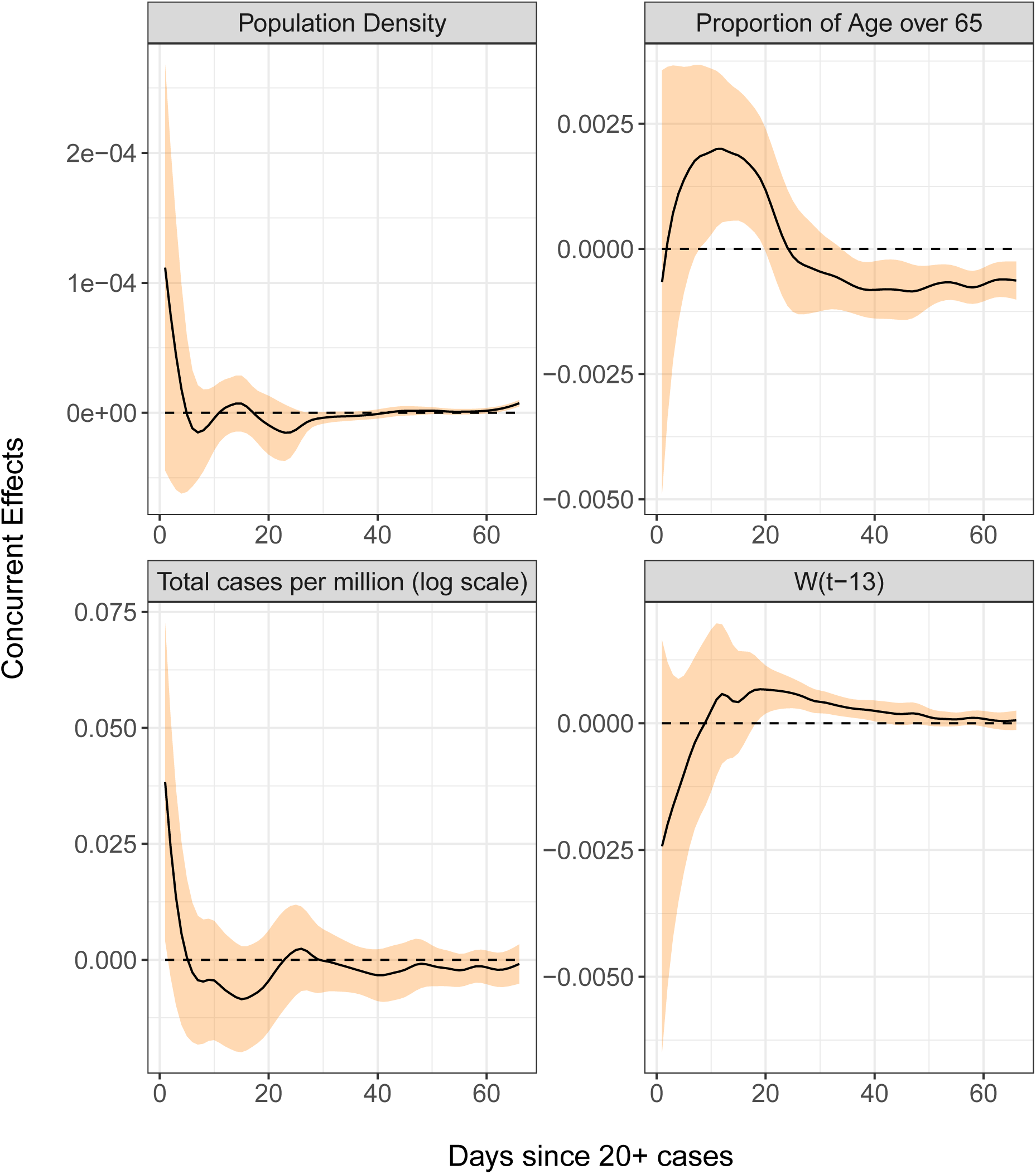
Time-varying regression effects of population density, the fraction of population over age 65, total cases per million and the percentage of the reduction from baseline in workplace mobility patterns thirteen days prior on doubling rate. The black curves are the concurrent effect functions and the orange ribbons represent 95% point-wise confidence bands. The variable *W*(*t* – 13) represents the percent change in workplace mobility lagged by 13 days.

##### Doubling Rates

Higher doubling rates reflect faster spread of infection and our analysis shows that they are not associated with higher population density in a consistent positive or negative way. The fraction of population over age 65 has a significant but complex effect on doubling rates. During the priming period, the doubling rate is positively correlated with demographically older populations, but this effect goes into reverse from day 25 on, perhaps as the presence of older members of society promotes additional self-isolation; however it is quite possible that this association is due to the potential confounding effect that many later mobility-reducing countries have older populations.

The additional effect of total cases per capita is significantly positive at the very start, but the concurrent regression slope estimate tends to be negative in the following period, which portends a dynamic regression to the mean effect [24], i.e., countries with doubling rates away from the mean tend to gravitate back to the mean during the early time period, reflecting declining variation across countries. The mean curve in Figure 6 indicates monotone declining doubling rates, so countries which did not have many cases in this period tend to catch up to the mean behaviour. Doubling rates are for the most part positively correlated with the size of the decline in workplace mobility with a lag of 13 days, which suggests that reduced mobility may significantly slow the growth rate of confirmed cases, though this benefit is only seen approximately 2 weeks later.

Figure 8 shows the predicted doubling rate for sixteen countries according to the historical functional linear model (see Section S.4 in the Supplement). Japan only has predictions after day 15 because the earlier mobility information is not available. Domains where the observed doubling rate is higher than the predicted indicate periods when the countries are doing worse than expected. The doubling rate of most countries is less predictable in the early stage but can be very well predicted later on, which may be partly a consequence of the dynamic regression to the mean effect as visualized in the left panel of Figure 6. Countries with relatively poorly predicted trajectories include Japan, India, Egypt, the Philippines and the US. This may be due to low testing rates, where the testing rate in Japan is reported to be much lower than other countries. The US had an incredibly low doubling rate in the beginning but also low levels of testing. It did much worse after 10 days and observations were in line with predictions only after day 35. Poland also behaved worse in the beginning but the doubling rate dropped to the predicted rate at day 10. The time points when improvement set in were about 1.5 to 2 weeks after the workplace mobility began to drop. This possibly indicates the lockdown policy was particularly effective in these two countries.

**Figure 8:**
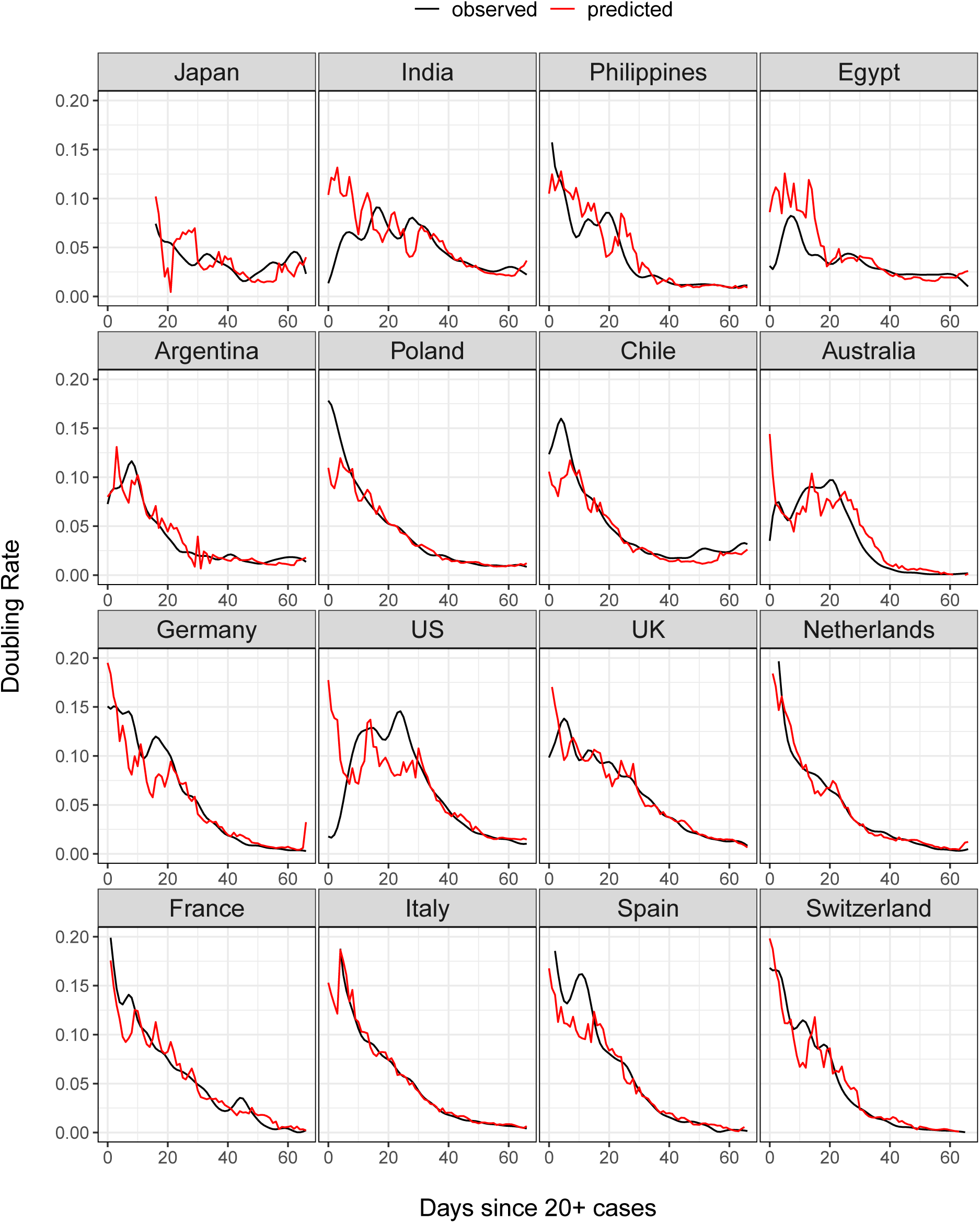
Observed and predicted trajectories for doubling rates of COVID-19 in sixteen countries, where predictors are total cases on log scale and workplace mobility with a 13 days lag. Decrease in workplace mobility is as presented in Fig. 5.

##### Case Fatality Rates

Case fatality rates exhibit explosive behavior dynamics as seen in the effect curve of *CFR*(*t*), which is positive after day 15. This means that case fatality rates either above or below the mean fatality rate curve in Figure 6 tend to move even further away from the mean as time progresses, as seen in Italy’s curve, for example. A possible explanation for this could be that an already overwhelmed healthcare system is predestined for future worse outcomes as the disease continues to spread and resources get even more scarce. Higher case fatality rates generally do not correlate strongly with population density, but are positively associated with demographically older countries, which is not surprising.

## 3 Discussion

In this study, we have explored a battery of functional approaches for modeling the cumulative COVID-19 case trajectories by pooling data across countries, which is facilitated by the curve-based methodology of functional data analysis. These techniques provide a natural and intuitive framework for comparing case trajectories, constructing principled forecasts for future case counts, and quantifying the time-varying effects of covariates. A major strength of the functional data approach is the borrowing of information across curves, i.e., models and predictions for case counts carry knowledge gained from the entire sample of curves and not just that of a single country’s data. This approach enables the modeling of time-varying associations and results in dynamic effects which are reflective of the changing nature of the infectivity cycles and their dependence on covariates. It also offers a flexible and non-parametric alternative to rigidly-structured epidemiological models.

The advantages afforded by a functionally-minded approach are not immune to limitations imposed by data quality, however. The bias in COVID-19 case count is difficult to control for, as the causes of under-reporting are several and complex (e.g., the delay between infection and confirmation, confounds in testing availability, prevalence of asymptomatic carriers, etc.). Findings regarding individual countries must be evaluated in light of the their unique set of biases. Indeed, this issue does not yet have a clear solution and warrants continued attention. A feature of our analysis that is favorable is that while absolute case counts may be subject to reporting error, the trends in time-dynamics on which we focus here are more robust, in the sense that biases for a given country may affect the entire trajectory, although to a different degree over time as for example testing is ramped up.

Applying functional PCA led to the discovery of four main patterns of disease progression since initial exposure and to identify the countries which follow them. A complementary analysis of percentlle rank dynamics allows for comparison of relative performance of countries at different points in time. In terms of predictive modeling, we introduce a dynamic FPCA approach for forecasting country-specific case counts, illustrated with 10-day forecasts. These forecasts, when compared to the eventually observed curves, can illustrate whether a country over- or under-performs during a given shorter time period. Lastly, we apply concurrent regression to quantify the effects of baseline and time-varying covariates, such as population density and reduction in social mobility over time. We distill the results of our analysis into seven key messages, summarized with their associated methods as follows:

- The mean trajectory of log-cumulative case counts per capita increases linearly (representing exponential growth) until around day 25 at which time it starts to bend. Variations from this mean trend behave according to a combination of two distinct and uncorrelated patterns of growth. The first pattern corresponds to a baseline level of viral spread which changes little over time; the second corresponds to a marked increase or decrease of the transmission rate after the first 25 days of exposure (Functional PCA, Figures 2 and 3).
- A country’s relative performance in terms of prevention stabilizes after an approximately month-long priming period. How well a country handled the situation during this initial window largely determined their final standing after 67 days of exposure (Rank Dynamics, Figure 4).
- Countries which were late to reduce social activity perform worse both relative to other countries and in terms of their own projected case counts (Dynamic FPCA, Figure 5).
- Trajectories of countries with earlier exposure times contain valuable information for forecasting case counts for more recently exposed nations. The curve-based methods of functional data analysis are uniquely suited for pooling information across a sample of trajectories, including those which are only partially observed. (Dynamic FPCA, Figure 5)
- Reduction in workplace mobility is associated with lower spread of the virus, though the effects are lagged. Benefits of reduced social mobility are delayed by roughly two weeks (Functional Concurrent Regression).
- Baseline demographics are significantly associated with doubling rates during specific time windows. More densely populated countries had higher initial rates of viral transmission, though this association faded after 20 days of exposure. Demographically older countries typically experience higher rates of spread during the first 25 days of exposure before course-correcting and enjoying lower doubling rates thereafter (FCR, Figures 6, 7 and 8).
- Case fatality rates exhibit positive-feedback patterns of severity. After the first two weeks of exposure, higher than average case fatality rates foreshadow increased mortality in the future. Meanwhile, countries which boast lower than average case fatality rates typically improve upon them even further. Demographically older countries also suffer higher case fatality rates, as expected. (FCR, Figures 6 and 9)

**Figure 9:**
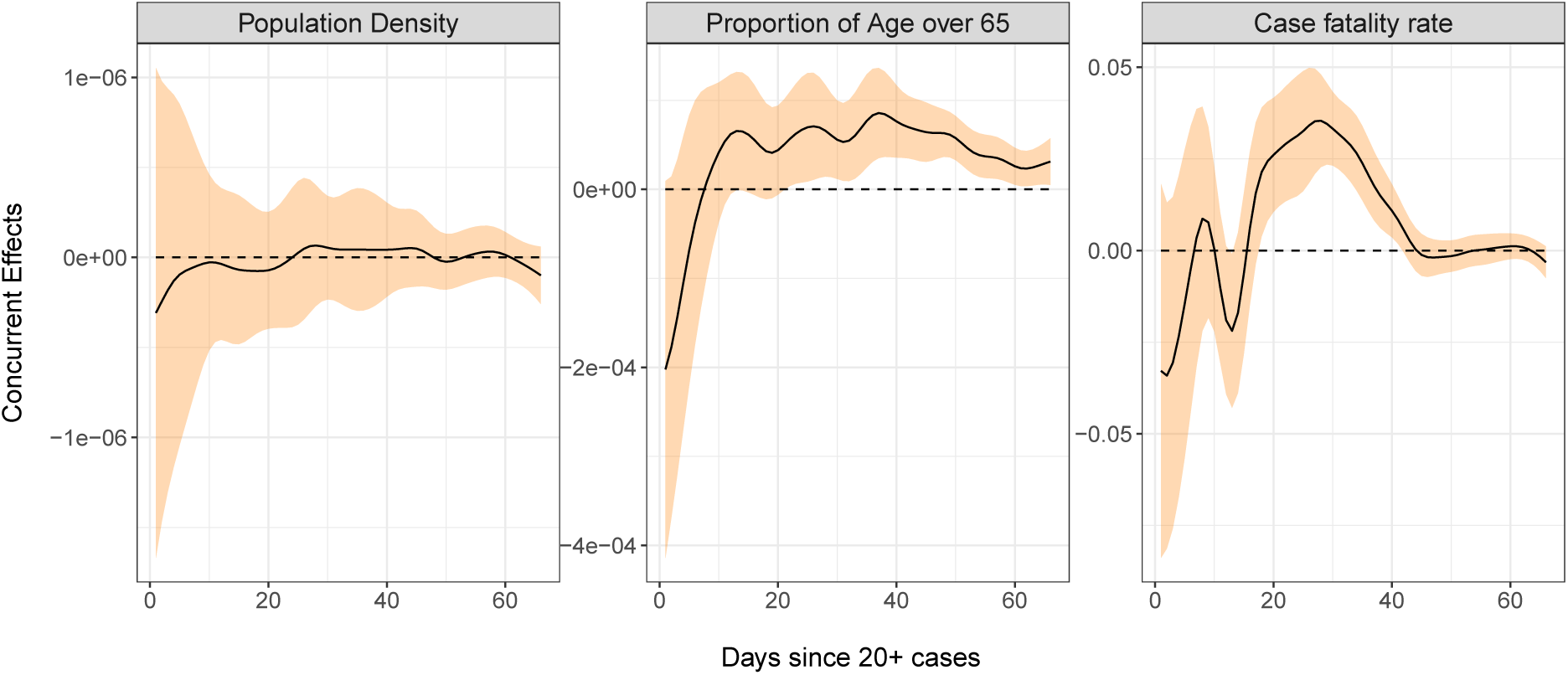
Time-varying regression effects of population density, percentage of population over age 65, and case fatality rates on the derivative of case fatality rate. The black curves are the regression slopes and the orange ribbon represents the 95% point-wise confidence band.

The first three key messages highlight the importance of early responses to the exposure, particularly during the first 25 days, a period that we have reason to characterize as a priming period. For example, the UK and Greece both have average baseline levels of spread, but Greece successfully maintains a level of containment while the UK suffers increased spread of the virus. The differences in initial responses across countries with similar baselines may be the key to understanding the factors which cause a country’s case load to escalate rapidly. The two patterns of growth identified here may serve as a useful jumping-off point for answering other similar lines of questioning. The importance of enacting preventative measures early is reaffirmed by our analysis, as countries who decreased social mobility before reaching high levels of spread did better than their predicted case counts and generally had less severe situations at the end of the interval.

The last three points highlight how factors may be relevant only during specific time periods. For example, one might hypothesize that a country with high population density would experience elevated transmission rates uniformly through time, as the population density is more or less constant. Our findings, however, suggest that this is not the case, as population density is uncorrelated with the doubling rate after the initial priming period. Careful consideration of baseline predictors with dynamic effects may indirectly reveal the mechanism through which the predictor acts on the response. Here, the detrimental impacts of high density living may be lessened or completely eliminated by the adoption of mobility reduction efforts.

The positive-feedback behavior seen in case fatality rates likely reflects the range in the ability of healthcare systems to cope with surges of cases. Struggling countries are expected to have elevated mortality rates, and as outbreaks become more severe, hospitals which are already overwhelmed suffer even higher rates of death. Conversely, we expect countries who have ample healthcare resources to have lower case fatality rates. Systems which are managing well eventually go on to enjoy even lower rates of mortality, perhaps as medical workers gain experience at treating the disease.

The code for implementing the methods employed for this analysis and also various other functional methods is publicly available in the R package fdapace [34].

## 4 Methods

### 4.1 Data

Our analysis focuses on modeling and predicting the cumulative number of cases per million individuals in log scale, using information obtained from the COVID-19 Data Repository by the Center for Systems Science and Engineering (CSSE) at Johns Hopkins University, which was accessed on May 18, 2020. The data consist of the cumulative number of confirmed cases and deaths per day since Jan 22, 2020 for several countries and is publicly available on Github at https://github.com/CSSEGISandData/COVID-19. Since the pandemic reached individual countries at staggered times, we consider a time interval 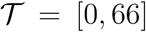 consisting of 67 days where the initial time *t* = 0 represents the earliest date at which at least 20 confirmed cases were reported.

The cumulative number of cases per million individuals on log scale is formally defined as

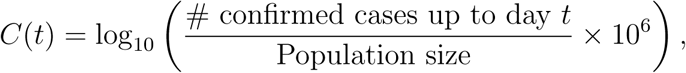

where *t* ∈ [0,66]. Defining the initial time *t* = 0 in such a way aligns trajectories according to the onset of the pandemic in a given country and allows for temporal comparisons across nations even if they were first exposed to the virus at different times. We include in our analysis countries with population size at least 100,000 by 2018 that have suffered at least 5 deaths by May 18, 2020 and have been exposed to the virus for at least 67 days, which results in *n* = 64 countries. A table of the selected countries and of the dates corresponding to *t* = 0 for all countries can be found in the Supplement S.1.

Data for population density and demographic percentage above 65 years old as of 2018 was obtained from the World Bank database which is available at https://www.worldbank.org/. For Iran we used the 2017 data as the 2018 input from worldbank was unavailable. Taiwan covariate information was not available in world-bank and was obtained from https://www.indexmundi.com/taiwan/#Demographics. We also use Google community mobility data as a time-varying covariate, available at https://www.google.com/covid19/mobility/. The following countries did not have Google mobility data available: Albania, Algeria, China, Iceland, Iran, and Russia. See Supplement S.1 for more details.

### 4.2 Functional PCA

The key to FPCA comes from the functional analogue of the spectral decomposition for covariance matrices. To extend this notion to functions, we consider a generic square-integrable stochastic process *C*(*t*), 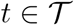 with mean function *μ*(*t*) = *E*(*C*(*t*)) and covariance function *G*(*s*, *t*) = Cov (*C*(*s*), *C*(*t*)), 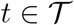.

Under certain conditions, *G*(*s*, *t*) admits an orthogonal expansion

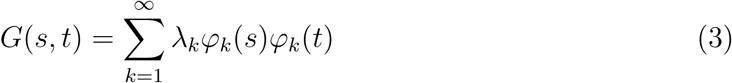

where λ_1_ ≥ λ_2_ ≥ ··· > 0, 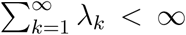, are the eigenvalues and *φ*_1_, *φ*_2_… are the (orthonormal) eigenfunctions of the Hilbert-Schmidt autocovariance operator 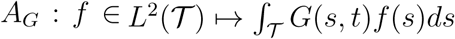.

With this decomposition, the Karhunen-Loève representation theorem states that

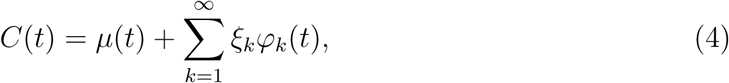

where the scores 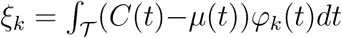 satisfy *E*(ξ*_k_*) = 0, Var(ξ*_k_*) = λ*_k_* and *E*(ξ*_k_*ξ*_i_*) = for *k* ≠ l. Here ξ*_k_* is the functional principal component score (FPC) of X(·) associated with the *k^th^* eigenfunction *φ_k_*. Thus, the FPC scores are projections of the centered stochastic process onto the directions given by the eigenfunctions and summarize how a function changes from the mean curve along the principal modes of variations. Moreover, from (4) the centered process is equivalent to ξ_1_, ξ_2_,….

By truncating the representation in (4) to a finite number of *K* components one achieves dimension reduction and can approximate the original stochastic process through its most important modes of variations. That is, FPCA provides a fit of the original curve

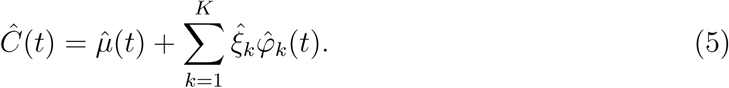

Here *K* is often chosen so that the fraction of variability explained, 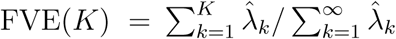, is above a threshold, e.g. 97%. For the COVID-19 cases, the choice *K* = 2 meets this criterion. Further details can be found in [29].

The modes of variation illustrate the individual effect that each of the scores has on the function *C*(*t*). The first mode of variation corresponds to the curve 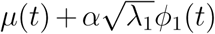, where *α* ranges in the interval [−2, 2]. This represents the effect of the first score on *C*(*t*) as it varies between ±2 standard deviations away from the mean. Similarly, the second mode of variation is obtained by replacing *λ*_1_ and *φ*_1_(*t*) by *λ*_2_ and *φ*_2_(*t*), respectively, in the previous expression.

### 4.3 Dynamic FPCA

The approximation in (5) provides a straightforward method for predicting a curve *C_i_*(*t*), if we have access to estimates of the mean function, eigenfunctions, and the FPC scores. We follow the Principal Components Analysis through Conditional Expectation (PACE) approach proposed for sparse longitudinal data [29]. For dynamic forecasting we proceed as follows: Suppose there are *n* countries for which trajectories have been observed for *t*_0_ + Δ*t* days, i.e., for the *i^th^* country, *i* =1, …,*n*, the current span of observations [0,*T_i_*] is such that *T_i_* > *t*_0_ + Δ*t*. If the number of countries *n* is not too small, the idea is to predict the future trajectory of the *n* + 1*^st^* country by borrowing information from the other n countries which have been observed for the time span to be predicted. Suppose we have data for the *n* + 1*^st^* country until time to *t*_0_ after the first 20 cases were reported but not beyond and are interested in predicting its trajectory for the next Δ*t* days, i.e., prediction in the time interval [*t*_0_, *t*_0_ + Δ*t*], where Δ*t* is chosen to be reasonably small. That is, we have observed data *C_n_*_+1_ = (*C_n_*_+1_(0),…, *C_n_*_+1_(*t*_0_))*^T^*.

One can then recover the process 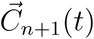 on [0, *t*_0_ + Δ*t*] by using {*C_i_*(*t_ij_*)}, where *i* = 1,…, *n* and 0 ≤ *t_ij_* ≤ *t*_0_ + Δ*t*. That is to say the forecast is obtained by using observations between day 0 to *t*_0_ + Δ*t* from other countries for which trajectories {*C_i_*(*t_ij_*)} have been observed for least for *t*_0_ + Δ*t* days. Then the estimated Karhunen-Loève representation (5) of *C_n_*_+1_(*t*) on the interval [0, *t*_0_ + Δ*t*] corresponds to

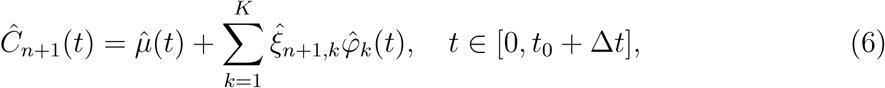

where 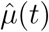 is estimated either cross-sectionally or with a smoothing method, 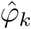 and 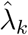 are estimated using the truncated spectral decomposition of the covariance surface, and the scores are estimated using the PACE approach [29], where the *k^th^* score for trajectory (*n* + 1) is

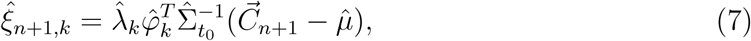

where 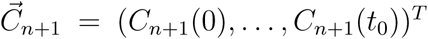 is the observed vector of data for the trajectory to be predicted, 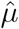 is estimated mean vector which equals to 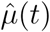 evaluated at *t* = (0, …, *t*_0_)*^T^*, and 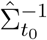 is the (*t*_0_ + 1) × (*t*_0_ + 1) estimated variance-covariance matrix for 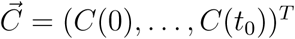 based on the entire sample.

We then iterate this process for each country of interest, and update our prediction moving forward. For forecasting COVID-19 cases, we display results for predicting Δ*t* = 10 days at a time.

## 4.4 Empirical Dynamics

The severity of infectious disease spread is commonly modeled using doubling time and case fatality rates. Here we describe time-varying regression models for these quantities inspired by empirical dynamics [24], which is an approach which systematically models the derivatives of smooth processes using the process itself. Technical details about empirical dynamics can be found in Section S.5 of the Supplement.

### 4.4.1 Doubling Rates

Let *N_c_*(*t*) denote the total confirmed cases at time *t*. Then the doubling time *κ*(*t*) at time *t* is such that

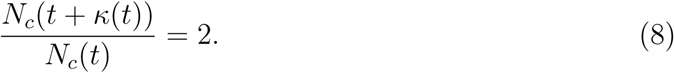

A Taylor series expansion leads to the approximation

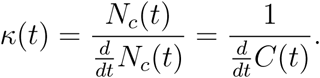

The *doubling rate γ*(*t*) is defined as

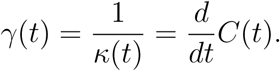

This naturally leads us to model the empirical dynamics of the process *C*(*t*) by

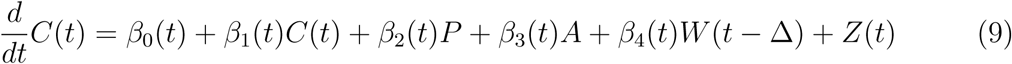

for *t* > Δ where *P* represents population density, A represents the proportion of population over age 65, *W*(*t* – Δ) denotes the percentage change from baseline of workplace mobility patterns at time *t* – Δ, and *Z*(*t*) denotes a mean zero stochastic drift process. We use the data adaptive criterion described in Section S.5 of the Supplement to select an optimal choice of lag Δ, which for our analysis is 14 days. Figures 3 and 4 in Section S.7 of the Supplement illustrate the strong multi-collinearity among the different community mobility patterns, so we utilize only workplace mobility. Note that in (9) we model the doubling rate *γ*(*t*) instead of the doubling time *κ*(*t*) which circumvents the numerical issues involved with a vanishing 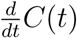.

For doubling rate prediction purposes we consider the functional linear regression model [35] that uses the entire history from *t* − 13 to *t* − 1 days,

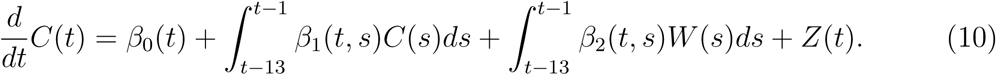

The details of this model are provided in Section S.5 of the Supplement.

### 4.4.2 Case Fatality Rates

The case fatality rate at time *t* is the ratio between the total death count and the total case count at that time. Letting *N_d_*(*t*) denote the total death count at time t, the case fatality rate is 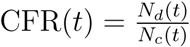. For the dynamics of fatality rates, we consider

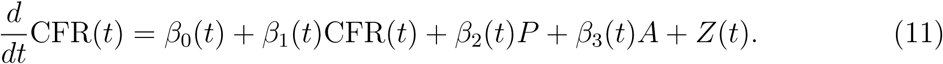

## 5 Data availability

Data that support the findings of this study are publicly available from the following sources: Johns Hopkins Univeristy CSSE (https://github.com/CSSEGISandData/COVID-19), World Bank (https://www.worldbank.org/), Indux Mundi (https://www.indexmundi.com/taiwan/#Demographics), and Google(https://www.google.com/covid19/mobility/). Scripts are publicly available in the R package fdapace (https://cran.r-project.org/web/packages/fdapace/index.html).

## Data Availability

Data that support the findings of this study are publicly available from the following sources:
Johns Hopkins Univeristy CSSE, World Bank, Indux Mundi, and Google. Scripts are publicly available in the R package fdapace.

https://github.com/CSSEGISandData/COVID-19

https://www.worldbank.org/

https://www.indexmundi.com/taiwan/#Demographics

https://www.google.com/covid19/mobility/

https://cran.r-project.org/web/packages/fdapace/index.html

**Author Contributions**
CC: Original draft, data analysis and interpretation, project administration
SB: Data analysis and interpretation
YC: Data analysis and interpretation
PD: Data analysis and interpretation
JF: Data analysis and interpretation
AG: Data analysis, acquisition, and interpretation
XZ: Data analysis and interpretation
JLW: Project administration and supervision, editing
HGM: Project conception, administration, and supervision, editing

